# Optimizing and Validating Systemic DNA Damage Response Profiling to Predict Neoadjuvant Chemoradiation Response in Rectal Cancer

**DOI:** 10.1101/2024.11.22.24317789

**Authors:** Elena V. Demidova, Philip Czyzewicz, Adria Hasan, Vladimir Avkshtol, Randy W. Lesh, Elizabeth Handorf, Karthik Devarajan, Bryant M. Schultz, Johanna D. James, Denise C. Connolly, Margret B. Einarson, Donald Baldwin, Erica A. Golemis, Joshua E. Meyer, Sanjeevani Arora

## Abstract

**Purpose:** This study aimed to stratify patients with locally advanced rectal cancer (LARC) based on their response to neoadjuvant chemoradiation therapy (nCRT) using DNA damage response (DDR)-related proteins measured in peripheral blood monocytes (PBMCs). We optimized and validated an innovative assay to quantify these proteins, providing a predictive framework for nCRT response.

**Experimental Design:** We used PBMCs collected from LARC patients either before or after standard course of ∼5.5 weeks of nCRT, with patients categorized by neoadjuvant rectal (NAR) score. DDR was assessed by immunofluorescence (γH2AX^S139^ foci), and by Luminex multi-analyte platform (xMAP) assay providing semi-quantitative assessment of phosphorylated Chk1^S345^, Chk2^T68^, γH2AX^S139^, p53^S15^ and total ATR, MDM2, p21. Assay performance was evaluated using reference controls and banked PBMCs from healthy controls (n=50).

**Results:** PBMCs from poor responders (PoR; NAR >14; n=21) had significantly lower γH2AX^S139^ foci than complete responders (CR; NAR <1; n=21) (p<0.0001), with no significant differences between pre- and post-nCRT samples (p=0.4961). The xMAP assay performance assessment showed linear sample curves, precision with acceptable inter- and intra-assay coefficients of variability, and high reproducibility with ∼1% outliers in replicates. Clinical associations using the xMAP assay found levels of six proteins (ATR, MDM2, Chk1^S345^, Chk2^T68^, γH2AX^S139^, p53^S15^) significantly differentiating CRs from PoRs (p ≤ 1e-5). Univariate CART analysis determined thresholds that segregated PoRs from CRs with high precision (p<0.001).

**Conclusion:** We optimized an assay to assess DDR proteins in PBMCs and identified specific proteins, along with their threshold levels, that can accurately predict response to nCRT in patients with LARC.

**Translational Relevance.:** Although neoadjuvant chemoradiation therapy followed by surgery is the standard of care for patients with locally advanced rectal cancer (LARC), many patients do not benefit from this treatment and suffer from its side effects. The motivation for this study was to reliably identify patients with LARC who will or will not respond to treatment, thereby permitting more effective direction of therapy only to likely responders. In this report, we describe identification and optimization of a novel multianalyte assay for patients diagnosed with LARC. This assay uses a Luminex xMAP platform to detect DNA damage response (DDR) signaling proteins in peripheral blood monocytes of pre-treatment patients. This assay, detecting the DDR proteins, effectively segregates responders from non-responders (p ≤ 1e-5), supporting optimization of treatment efficacy and reduction of unnecessary toxicity, thus advancing personalized medicine in oncology.

## Introduction

Neoadjuvant chemoradiation therapy (nCRT) followed by surgery is a standard of care for patients with locally advanced rectal cancer (LARC) (clinical stage II-III) (1,2), accounting for ∼60% of newly diagnosed rectal cancer cases (3). nCRT is effective in improving locoregional control for many patients (4). Approximately 20% of patients undergoing nCRT achieve a pathologic complete response (pCR), while around 25% show limited or no response (4–8). Attempts to predict nCRT response through anatomical imaging (9,10) or tumor sequencing (11) have been unsuccessful, and current clinical and radiographic factors are inadequate in identifying which patients will benefit most from nCRT (12–15). Consequently, many patients endure the toxic side effects of nCRT without deriving significant therapeutic benefit. A reliable strategy to predict nCRT response could greatly enhance personalized treatment for rectal cancer.

The primary therapeutic mechanism of radiation-induced cell death is through induction of DNA damage in tumors at levels that cannot be repaired by the cellular DNA repair machinery (16,17). Radiotherapy induced DNA damage including double stranded breaks (DSBs), DNA cross links, base damage, and single strand breaks (18). Combining chemotherapy with radiation, particularly 5-fluorouracil-based nCRT, further amplifies DNA damage by inhibiting key enzymes involved in DNA synthesis, such as thymidylate synthase (17,19,20).

While DNA repair plays a crucial role in chemotherapy and radiation responses, no tumor-based gene or protein signatures related to DNA repair have led to effective predictive biomarkers for nCRT outcomes in LARC. Studies have explored various biomarkers, including tumor mutations, gene expression profiles, and tumor microenvironment infiltrates (21–24), but none have proven predictive. This is an important therapeutic gap, as radiation also impacts adjacent healthy tissues, leading to side effects such as radiation-induced diarrhea, rectal bleeding, and dermatitis (25–27). Reliably identifying likely responders versus non-responders would reduce the incidence of these sometimes-serious adverse events.

To address this critical challenge, we hypothesized that the innate DDR capacity of peripheral blood monocytes (PBMCs) could serve as novel predictive tool for nCRT response in LARC. PBMCs offer a minimally invasive alternative to tumor biopsies and provide insight into systemic DDR capacity, which could better reflect a patient’s ability to respond to DNA-damaging therapies. Germline alterations in DNA repair genes are known to affect patient responses to such therapies (28–34), and our prior work has demonstrated that defects in DNA repair can be detected using assays of DDR proteins, such as γH2AX, in circulating lymphocytes (35–37).

In this study, we hypothesized that the pre-treatment DDR capacity of PBMCs could predict response to nCRT in LARC patients. By focusing on circulating immune cells rather than tumor tissue, we aim to overcome the limitations of tumor-based markers and offer a new method for stratifying patients based on their systemic DDR capability. Here, we optimized and validated a novel multi-analyte assay, identifying six proteins that effectively segregate patients based on their therapeutic response. This innovative, blood-based assay presents a promising tool for enhancing personalized treatment strategies, potentially improving both treatment outcomes and quality of life for LARC patients.

## Material and methods

### Patient population

The work was approved by the FCCC (Fox Chase Cancer Center) Institutional Review Board (IRB 18-4005 and 22-9925). All patients were diagnosed with non-metastatic, locally advanced rectal adenocarcinoma (T3/4 or node-positive) and treated with nCRT. Radiation treatment consisted of 50-50.4 Gy of external beam radiation therapy delivered in 25 to 28 fractions over 5 to 5.5 weeks. Patients received concurrent 5-fluorouracil delivered via continuous infusion (225 mg/m^2^) or oral capecitabine (825 mg/m^2^ twice daily) on days of radiation. Surgical resection was performed ∼8-10 weeks after nCRT completion. Patients were selected for analysis to provide a distribution of nCRT responses and NeoAdjuvant Rectal (NAR) scores. The patient samples analyzed in this study (viably stored PBMCs: pre-nCRT [treatment naïve], post-nCRT [∼8-10 weeks], DNA, and serum) were obtained from the FCCC Biosample Repository Facility (BRF) and Department of Radiation Oncology. Samples were collected under consent for research and deidentified or coded.

### PBMC banking, T-cell culture, and immunofluorescence imaging

Experiments analyzing γH2AX^S139^ foci in PBMCs as an indicator of DSBs were performed as previously described (35,36). PBMCs were fractionated from patients’ blood and preserved by the FCCC BRF using standard methods to maintain viability. To induce proliferation, PBMCs were stimulated with phytohemaglutinin (PHA), M form (Life Technologies, Carlsbad, CA) at a final concentration of 1.5%, and 50 units/ml recombinant human interleukin 2 (IL-2) (NCI Preclinical Repository) (35). Cells were left untreated (baseline) or treated with 2.0 Gy IR, and then incubated for 1h. The cells were either fixed in paraformaldehyde, and immunofluorescence analysis was performed using an antibody to γH2AX^S139^ (clone JBW301, MilliporeSigma Inc.) and DAPI (Life Technologies) to stain the nucleus. The γH2AX^S139^ foci were quantified using the ImageXpress micro-automated high-throughput microscope (Molecular Devices, San Jose, CA) driven by MetaXpress software, as previously described (35,36).

### Viability of EBV-transformed PBMC cell lines

PBMCs from LARC patients were collected and immortalized with Epstein-Barr virus (EBV) as previously described (35,36). Briefly, lymphocytes isolated from 10 ml blood by centrifugation over Ficoll-Paque were resuspended in 2.5 ml complete RPMI-1640 without insulin. Immortalization was initiated by adding EBV strain B95-8 as 1.0 ml of filtered supernatant from the marmoset B-cell line GM 7404. After incubation for 2h at 37 °C, 6.5 ml of complete RPMI containing 1 mg/ml cyclosporine A (Sigma-Aldrich, CA) was added, and the suspension was cultured at 37 °C, 5% CO_2_ in an up-right flask. After 3 weeks, the culture was split, and 5 ml fresh medium was added to each flask. EBV-transformed PBMC cell line from LARC patients, both PoRs (n=6, NAR-scores: 20.4 & 14.98) & CRs (n=6, NAR-scores:0.94) were used. Cells were treated with different doses of irradiation (2 Gy to 50 Gy), oxaliplatin (0.25 µM to 75 µM; #25021-233-20, Sagent Pharmaceuticals, IL), irinotecan (0.5 µM to 150 µM; #25021-230-05, Sagent Pharmaceuticals, IL), or 5-fluorouracil (0.5 µM to 150 µM; #16729-276-38, Accord Healthcare, Durham, NC). Cell viability was measured by short-term CellTiter-Glo® assay (#G7570, Promega Corporation, Madison, WI) 72h after drug treatment.

### Luminex xMAP immunoassay

The assay was performed using the MILLIPLEX® DNA Damage/Genotoxicity magnetic bead panel kit (48-621MAG, Millipore Sigma, Burlington, MA). The 48-621MAG panel was used to quantitate the levels of the following proteins: total ATR, Chk1^S345^, Chk2^T68^, γH2AX^S139^, p53^S15^, total MDM2 and total p21. The immunoassay kit is provided with lyophilized MILLIPLEX MAP Cell Lysate (Catalog # 47-229, 47-207 and 47-218) controls that were used to create calibration curves for the 7 analytes in the assay. The BioPlex 200 platform (Bio-Rad, Hercules, CA) in the FCCC High Throughput Screening Facility (HTSF) was used to generate the xMAP data. The detailed immunoassay protocol is in the **Supplementary document**.

### Statistical Analysis

Patient characteristics, including demographic, tumor, clinical, and nCRT toxicity, were summarized. DDR proteins were assessed in all study patients. The foci or levels of DDR-related proteins was described graphically and using standard statistical methods. Relationships between DDR-related proteins were characterized using Pearson and Spearman’s correlations. Pre/post differences of proteins within patients were tested using Wilcoxon signed-rank tests for paired data (where both pre- and post-nCRT samples were available). For cell line viability, Mann-Whitney tests were used.

The % coefficient of variation (% CVs) were used to describe variation among replicates for all performance characteristics. Both inter- and intra-assay CVs were used for precision analysis. Data from all replicates were used for precision analysis. For intra-assay precision, the %CV was calculated for the duplicates on the plate. To determine inter-assay precision, all replicates across all plates were pooled together to determine the mean and the standard deviation (SD). The %CV were calculated for these data using the pooled mean and standard deviation values.

For assessments of reproducibility of data from the Luminex xMAP assay, replicate measurements of expression were obtained for each protein for lysates from banked and banked-cultured PBMCs. All negative values were truncated to zero and replicate pairs with either missing observations or identical values across duplicates were removed from further analysis as these were deemed to be non-informative for this analysis. The coefficient of variation (CV) and the difference (D) in duplicate observations for each replicate pair were used in detecting outlying pairs after accounting for inherent noise in the assay measurements. Kernel density plots and quantile-quantile (QQ) plots were used to empirically determine the distributions of these quantities, and maximum likelihood methods were used to estimate the parameters. Observations within one standard deviation of the mean of the empirical distribution for D (determined to be asymmetric Laplace) were considered to be within acceptable limits and excluded from further analysis. The middle 90% of the remainder of the data was then fit to the QQ plot positions of the empirical distribution of CV (determined to be log-normal). A test was performed to determine whether extreme observations are outliers by computing the top and bottom thresholds beyond which exactly a pre-specified number of observations are expected. Any observation that lies beyond these thresholds has a very low probability of being generated by a log-normal model. Observations that lay above the top threshold or below the bottom threshold *and* beyond one standard deviation from the mean of the empirical distribution of D were identified to be outliers. A number of choices for the pre-specified number of observations were considered in order to evaluate the robustness of this method to identifying outliers. More details on this approach can be found in (38). Computations were performed in the R statistical language and environment using libraries VGAM and extreme values (38–42).

Due to the hypothesis generating nature of this study, no adjustment for multiple hypothesis testing was done. All tests were two-sided and used a Type I Error of 0.05 to determine statistical significance unless specified otherwise. The classification and regression trees (CART) methodology was used to identify statistically significant cut-points that delineated patient response. This approach utilized the unified CART framework that embeds recursive binary partitioning into the theory of permutation tests (43) and implemented in the R package party (44).

### Data Availability Statement

The data generated in this study are available within the article and its supplementary data files. Additionally, raw data may be available upon request from the corresponding author.

## Results

### Elevated positive basal γH2AX^S139^ foci in PBMCs from LARC patients’ correlates with response to nCRT

We identified 54 patients with LARC (**Figure 1A**) who had varying responses to nCRT and thus a range of NAR scores (a measurement of response and predictor of overall survival in rectal cancer, (44)). The majority were male, with median age 55 years (range 25-82). Most (94%) had T3 priy tumors and N0 disease (72%) (**Table 1**, independent set 1). The identified patients were separated into nCRT response groups based on NAR score (**Figure 1A top**); response groups were designated as complete response (CR; NAR<1, n=21), partial response (PR; 1<NAR<14, n=12), and poor response (PoR; NAR>14, n=21). For each patient, PBMCs were collected either pre-treatment (pre-nCRT) or prior to surgery (pre-Surg after completion of nCRT).

**Figure 1.**
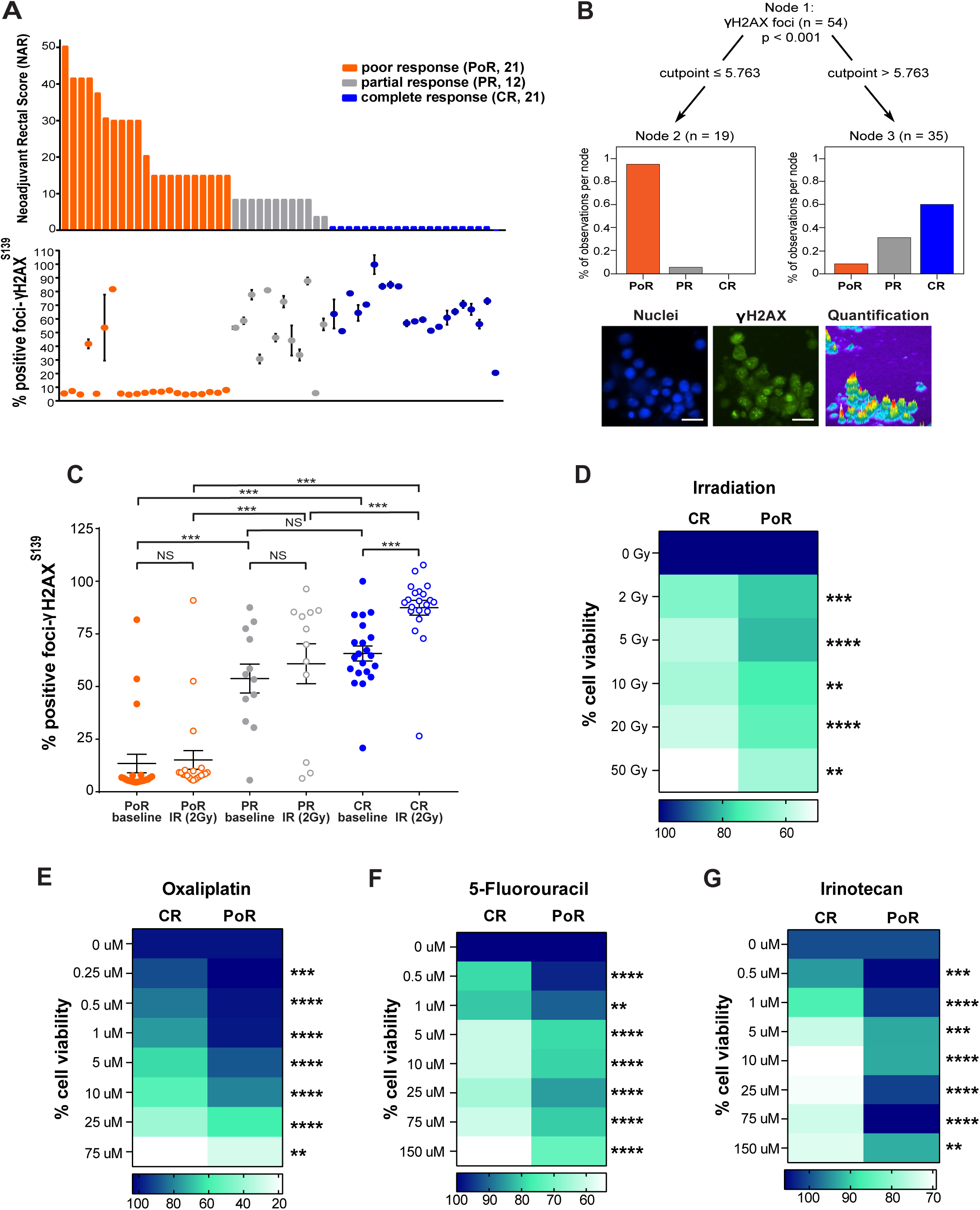
Increase in positive γH2AX^S139^ foci in PBMCs from LARC patients correlates with complete response. **(A)** Correlations of patient-level data with nCRT response. Top: nCRT response as assessed by NAR, with patients grouped into PoR (orange; n=21), PR (grey; n=12), CR (blue; n=21). Bottom: Comparison of γH2AX^S139^ foci results to the NAR score in PBMCs from 54 patients. Results are shown as% positive γH2AX^S139^ foci normalized to complete responders. Each filled circle represents a patient. Error bars indicate SEM (n=3). **(B)** Conditional inference tree for association of the Combined DDR Score with response. The 1^st^ node represents the total number of cases (n=54) with the branches that lead to specific binary partition based on the optimal cutpoints. The y-axis shows the percentage of response group per node. Bottom: Representative example of the high-throughput, automated, immunofluorescence-based, image acquisition used to identify and analyze γH2AX^S139^ foci. Primary PBMCs are labeled with an antibody for γH2AX^S139^. DAPI (blue) shows nucleus, γH2AX^S139^ foci (green). Foci are quantified for pixel intensity, size, and shape. Scale bar =20µm. **(C)** Quantified γH2AX^S139^ foci in PBMCs before and after IR. Banked PBMCs were cultured from all cases and were treated with IR (2 Gy), fixed, stained for γH2AX^S139^ foci, imaged and quantified. The data are presented as percent positive γH2AX^S139^ foci, normalized to highest raw value of positive γH2AX^S139^ foci. Each filled circle represents a patient pre-IR treatment; open circles represent patient post-IR treatment. **p < 0.01, ***p < 0.001, NS (non-significant), using Kruskal-Wallis tests with by pairwise Wilcoxon rank sum tests. Complete responder (CR)-patient derived cell lines have significantly lower cellular viability post DNA damage compared to poor responder (PoR)-patient derived cell lines. **(D-G)** PBMCs from CR had elevated γH2AX^S139^ foci after IR-treatment vs. PR and PoR. EBV-transformed lymphoblastoid cell lines from CRs (n=6) and PoRs (n=6) were either left untreated or treated with **(D)** irradiation, **(E)** oxaliplatin, **(F)** 5-Fluorouracil and (**G)** irinotecan at the doses indicated. Cell viability was measured by CellTiter-Glo® assay 72h after treatment. Aggregate data from the six individual cell lines under each response group are shown as % cellular viability (n=3). (IR, 2-50 Gy, p<0.01; oxaliplatin, 0.25-75 µM, p<0.01 for 75 µM, and p<0.001 for others; 5-FU, 0.5-150 µM, p<0.01 for 1 µM, and p<0.0001 for others; and irinotecan, 0.5-150 µM, p<0.01 for 150 µM, and p<0.001 for others). *p<0.05, **p<0.01, ***p<0.001, ****p<0.0001, NS - not significant. Mann-Whitney Test.

**Table 1.** Demographic and clinical characteristics of LARC patients in this study.

| Characteristic | Independent Set 1 | Independent Set 2 |
| --- | --- | --- |
| **Age (years)** |  |  |
| Mean | 56 | 60 |
| Median | 55 | 62 |
| Range | 25-82 | 31-82 |
| **Gender, total N (%)** |  |  |
| Male | 36 (67) | 28 (64) |
| Female | 18 (33) | 16 (36) |
| **Self-reported Race, total N (%)** |  |  |
| Black | 3 (6) | 6 (14) |
| White | 50 (92) | 38 (86) |
| Asian | 1 (2) | 0 (0) |
| **Pre-treatment clinical T classification, total N (%)** |  |  |
| 1 | 0 (0) | 0 (0) |
| 2 | 2 (4) | 1 (2) |
| 3 | 51 (94) | 38 (86) |
| 4 | 1 (2) | 5 (12) |
| **Post-treatment pathological T classification, total N (%)** |  |  |
| 0 | 20 (37) | 15 (34) |
| 1 | 3 (5) | 1 (2) |
| 2 | 11 (20) | 3 (7) |
| 3 | 14 (26) | 22 (50) |
| 4 | 3 (6) | 3 (7) |
| X | 3 (6) | 0 (0) |
| **Pre-treatment clinical N classification, total N (%)** |  |  |
| 0 | 39 (72) | 8 (18) |
| 1 | 13 (24) | 31 (70) |
| 2 | 1 (2) | 3 (7) |
| X | 1 (2) | 2 (5) |
| **Post-treatment pathological N classification, total N (%)** |  |  |
| 0 | 42 (77) | 27 (61) |
| 1 | 8 (15) | 10 (23) |
| 2 | 2 (4) | 6 (14) |
| X | 2 (4) | 1 (2) |

To initiate analysis of DDR levels as a correlate of response, PBMCs from each LARC patient (independent set 1) were cultured and analyzed for basal intensity of γH2AX^S139^ nuclear foci, a common marker of DSBs (35) (**Figure 1A bottom**). Here, no significant differences in positive γH2AX^S139^ foci were observed between PBMCs collected pre-nCRT and PBMCs collected pre-surgery after nCRT completion from the same patient (p=0.4961, n=11, **Supplementary Figure 1A**). This excluded nCRT-induced changes in PBMCs as an explanation for differences between individuals and suggested that data from both time points could be combined for further analysis. Number of positive γH2AX^S139^ foci were not related to subject age (**Supplementary Figure 1B**, Spearman’s correlation=-0.03, p=0.8315).

We then used the mean baseline positive γH2AX^S139^ foci scores to obtain an optimal cut point that could segregate the response groups. Each terminal node of the test data represents a subgroup of the full set created by partitioning at the chosen cut point (**Figure 1B**). Based on this procedure, a single cut point was identified which best separated the patients by response (**Figure 1B**, node 2 and 3). For PBMCs with a mean γH2AX^S139^ score ≤5.763, 95% belonged to the PoR group. For PBMCs with a mean γH2AX^S139^>5.768, 60% belonged to the CR group and ∼10% to the PoR group. **Figure 1B** (**bottom**) shows representative images for the primary PBMCs labeled with an antibody for γH2AX^S139^ (green) and nucleus labeled with DAPI (blue).

### NAR score correlates with positive γH2AX^S139^ foci induction in PBMCs in response to ionizing radiation (IR)

The relationship observed between positive γH2AX^S139^ foci and NAR could solely reflect static properties of patient cells *in vivo* or could reflect differences in dynamic capacity of patient cells to respond to DNA damage, or both. To evaluate the baseline and IR-induced response of PBMCs, γH2AX^S139^ foci in the PBMCs from the CR, PR and PoR groups of patients were visualized by IF and quantified at baseline or following treatment with 2.0 Gy IR (**Figure 1C**). When comparing these groups, there was a significant difference between the CR-IR group versus the PoR-IR and PR-IR groups. Interestingly, the PoR-IR and PR-IR groups did not show significant differences in γH2AX^S139^ foci post-IR, whereas the CR-IR group exhibited a significant *in vitro* response beyond the already robust intrinsic levels of γH2AX^S139^ foci.

To systematically examine the value of γH2AX^S139^ foci as predictive of response to additional DNA-damaging treatments, we generated LARC patient-derived cell lines from the primary PBMCs. Of 12 cell lines were derived from PBMCs, 6 were from PoRs and 6 were from CRs. We treated these patient-derived cell lines with different doses of IR (**Figure 1D**), oxaliplatin (**Figure 1E**), 5-FU (**Figure 1F**), and irinotecan (**Figure 1G**). Cellular viability between the response groups was assessed by short-term end point CellTiter-Glo® assay (72h after the drug treatment). Here, paralleling results with primary cells, significantly decreased cell viability was observed following radiation or drug treatment for cell lines from the CR group of patients compared to the cell lines from the PoR group of patients.

### Optimizing DDR xMAP assay for qualitative and quantitative assessment of DDRs

Measures of γH2AX^S139^ foci by IF can be inconsistent due to inter- and intra-lab variability(12,35,36,45–48). The ability to reliably quantitate γH2AX^S139^ would be highly beneficial in validating an assay for clinical settings. To this end, we optimized a Luminex Multi-Analyte Platform (xMAP) assay to measure DDRs (hereafter the assay is referred to as the DDR xMAP assay). The DDR xMAP assay not only detects γH2AX^S139^ but also simultaneously detects multiple other proteins involved in activated DDR signaling (Chk1^S345^, Chk2^T68^, p53^S15^, and total ATR, MDM2, and p21). To apply this assay to LARC patient PBMC biospecimens, we developed calibration curves for qualitative and quantitative assessment of DDR proteins.—**Figure 2A, 2B** and **Supplementary figure 2** show the semi-quantitative assessment of the above DDR proteins using assay controls. The calibration curves provide upper and lower limits of detection for each protein. The treatment specific assay controls confirm no cross-reactivity and that signal from each protein in the assay is not due to any non-specific interactions. **Figure 2C** shows high reproducibility of assay controls, with only 2 outliers identified by our approach from a total of 5688 replicate pairs of observations (i.e., 0.035% outliers). **Supplementary table 1** shows the degrees of freedom, R^2^ values, adjusted R^2^, and sum of squares obtained for the assay controls 1 and 2.

**Figure 2.**
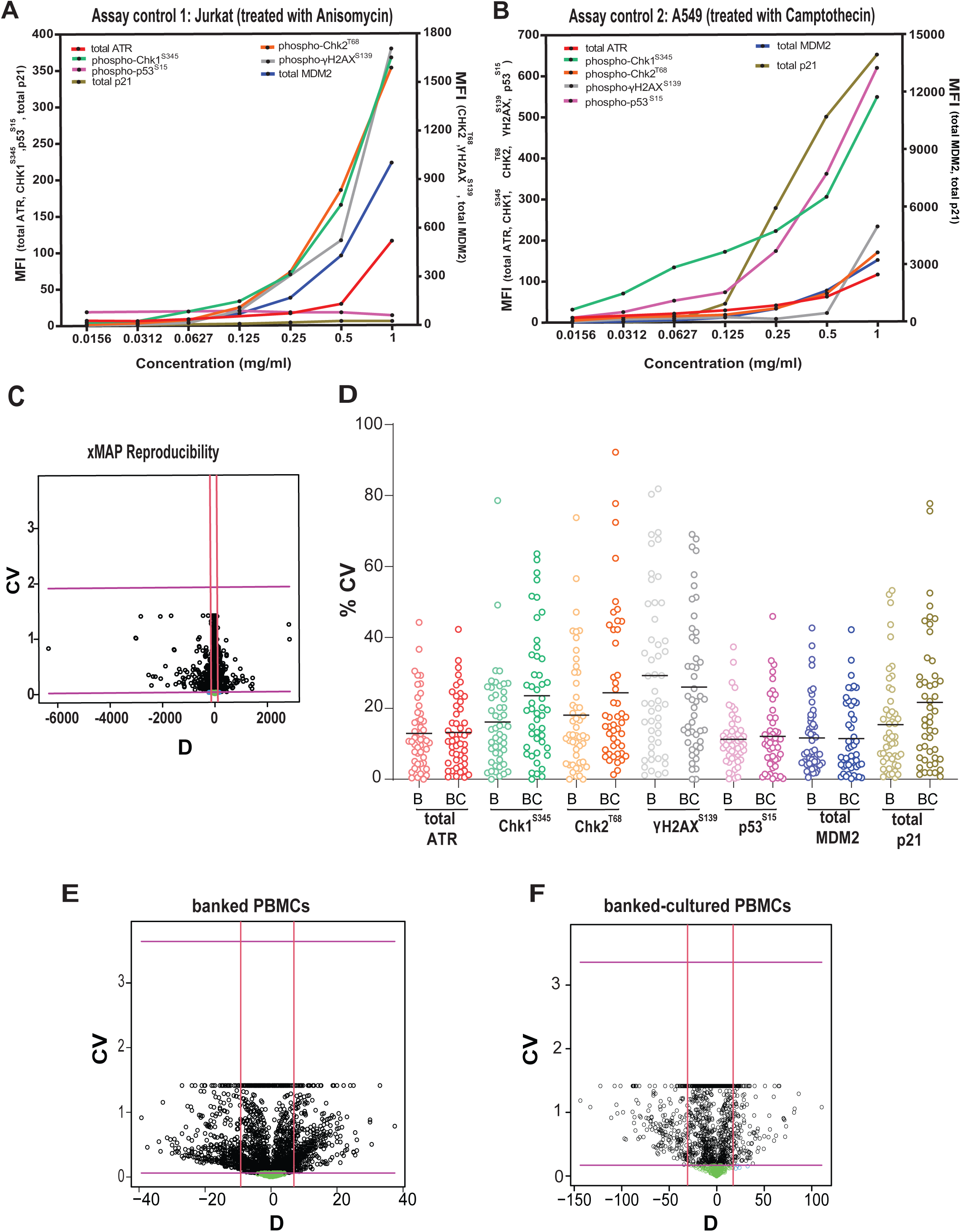
DDR xMAP optimization and validation. **(A, B)** Graphs show development of a semi-quantitative curve using vendor-provided assay controls for the seven DDR-related protein markers. The figure shows aggregate data for the dilution of assay control 1 (Jurkat treated with anisomycin) and assay control 2 (A549 treated with camptothecin) representing data from 12 replicates. The controls are differentially positive or negative based on the marker tested and serve as internal controls of assay performance. **(C)** A scatterplot of the CV, versus the difference, D, for all pairs of replicates is used to assess reproducibility of the data. The red vertical lines represent one (1) standard deviation (of D) away from its estimated mean and green circles within this band represent replicates exhibiting extreme values of CV (large or small) but with small D. The maroon horizontal lines represent the outlier determination thresholds for CV and replicates whose CV is above the level indicated by top horizontal line or below the level indicted by the bottom horizontal line and whose D is outside this band of red vertical lines are outliers (shown as blue circles). Replicates shown as green and black circles are not considered to be outliers. **(D)** Graphs show % CV comparison of the seven DDR-related protein markers in lysates prepared from banked (denoted as B) and banked-cultured PBMCs (denoted as BC). **(E, F)** A scatterplot of the CV versus the difference (D) showing no significant difference in lysates from banked vs banked-cultured PBMCs. The red vertical lines represent one (1) standard deviation (of D) away from its estimated mean and green circles within this band represent replicates exhibiting extreme values of CV (large or small) but with small D. The maroon horizontal lines represent the outlier determination thresholds for CV and replicates whose CV is above the level indicated by top horizontal line or below the level indicated by the bottom horizontal line and whose D is outside this band of red vertical lines are considered to be outliers (blue circles). Replicates shown as green and black circles are not considered to be outliers.

### DDR xMAP Assay Pre-analytical Validation

We next determined whether the DDR proteins measured by the DDR xMAP assay would remain stable in lysates prepared from banked PBMCs versus cultured PBMCs. Avoiding the step of culturing PBMCs is critical for the development of a clinically relevant assay. To assess this, we used PBMCs from the same healthy individuals (**Supplementary Table 2**) to compare data from two different lysate preparation methods: (1) lysates prepared directly from banked PBMCs without prior culturing (referred to as banked PBMCs), and (2) lysates prepared from banked PBMCs that were cultured prior to lysate preparation (referred to as banked-cultured PBMCs). **Figure 2D** shows percentage CVs for each protein (total ATR, Chk1^S345^, Chk2^T68^, γH2AX^S139^, p53^S15^, total MDM2 and total p21) when either banked PBMCs or banked-cultured PBMCs. In the analysis of data from the banked PBMCs, no outliers (out of 4320 replicate pairs) were identified (**Figure 2E**). However, a total of 4 outliers (out of 1800 replicate pairs or 0.22%) were identified in the data set representing banked-cultured PBMCs (**Figure 2F**). The scatterplots of % CV vs. difference (D) appear very similar for these two sets of data with one important distinction: the range of D for banked PBMCs (-40,40), is markedly narrower than that for banked-cultured PBMCs, (-150,110), despite a larger number of replicate pairs of observations available for banked PBMCs (4320 pairs from 6 replicates vs. 1800 pairs for banked-cultured PBMCs from 4 replicates). In addition to the absence of any outliers, this narrower range of differences in replicate pairs for banked PBMCs indicates better reproducibility overall compared to banked-cultured PBMCs. **Supplemental figures 3 and 4** display detailed graphs of aggregate data from lysates prepared from banked PBMCs and banked-cultured PBMCs from healthy controls, plotted along the calibration curves of each vendor-provided assay control.

### Application of the optimized DDR xMAP in LARC patient PBMCs and correlations with response to nCRT

We applied the validated DDR xMAP assay to PBMC lysates from two independent sets of LARC patients (**Table 1, independent set 1 and 2**). The second independent set of 44 LARC patients (**Table 1**, independent set 2) contained two response groups of nCRT (CR and PoRs). The majority were male, with median age 60 years (range 31-82). Most (86%) had T3 primary tumors and N1 disease (70%) (**Table 1**, independent set 2). For each patient, PBMCs were collected, either pre-treatment or pre-Surg (same as for set 1).

First, we analyzed PBMCs of LARC patients from the independent set that contained the three nCRT response groups (n=48, **Table 1**, subset of independent set 1 containing the CR, PR and PoR groups). When all response groups were included in the analysis, we found that at baseline the CR group had significantly elevated expression of total ATR, Chk1^S345^, and Chk2^T68^, activation of γH2AX^S139^, checkpoint-associated activation of p53^S15^, and total MDM2 (**Figure 3A-F**). Here, γH2AX^S139^ protein levels (**Fig 3D**) were elevated in the CR and PR groups compared to the PoR group, in agreement with the previous positive γH2AX^S139^ foci results (**Figure 1**). When comparing data between the CR and PR groups, levels of four individual proteins could each significantly differentiate them (total ATR, Chk2^T68^, p53^S15^ and total MDM2, in **Figure 3A,** 3C, 3E, and **3F**). When comparing the PR and PoR response groups, only γH2AX^S139^ protein levels could significantly differentiate them (**Figure 3D**). For some of the DDR-related proteins, including notably Chk1^S345^, the PR patients had a higher average value than the PoR group that neared significance (p=0.078) (**Figure 3B**). Importantly, levels of six individual proteins (total ATR, γH2AX^S139^, total MDM2, Chk1^S345^, Chk2^T68^ and p53^S15^) could significantly differentiate CR from PoR. Finally, levels of total p21 did not correlate with response status (**Figure 3G**).

**Figure 3.**
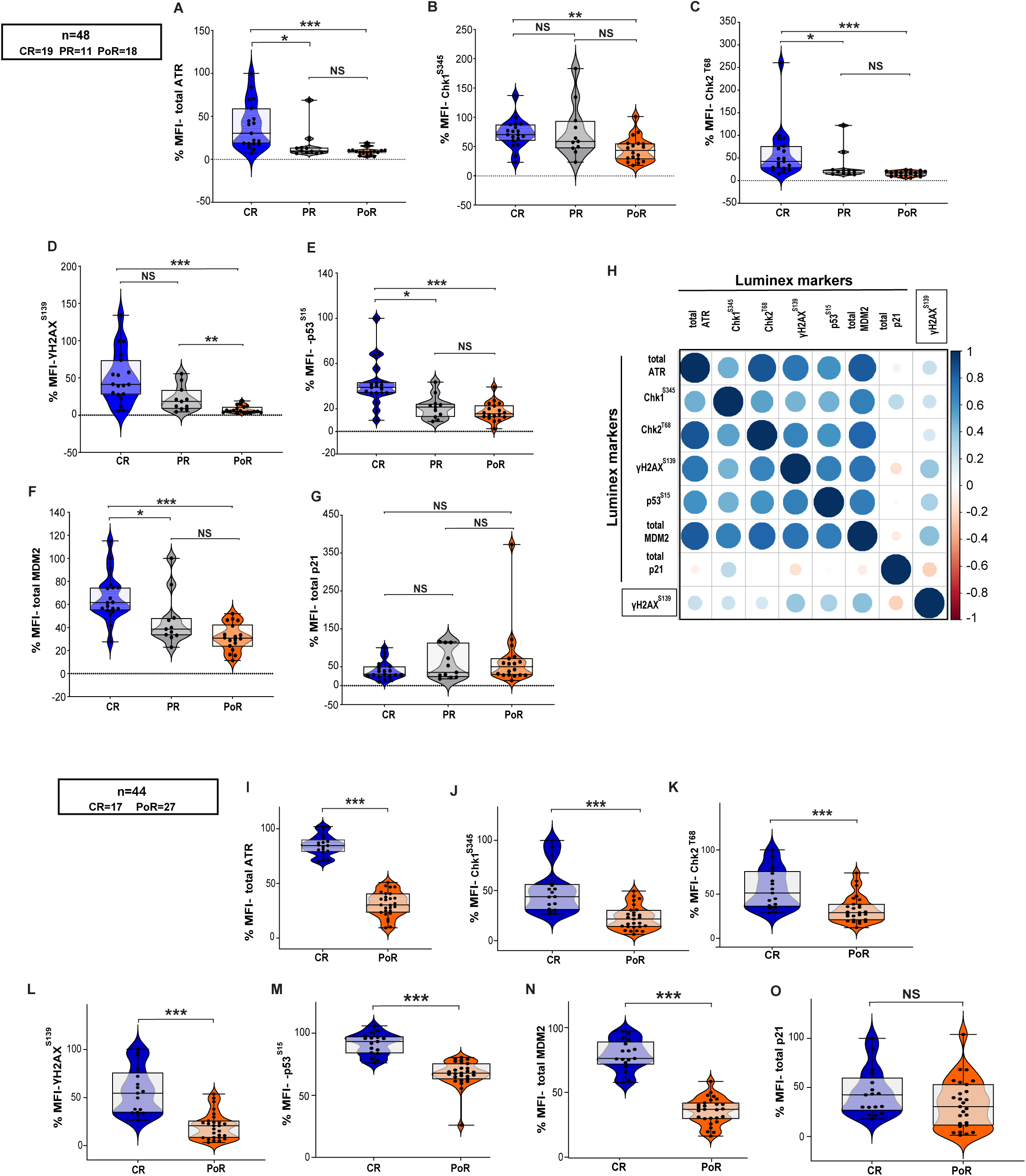
Association of the seven DDR-related protein markers in PBMCs of LARC patients with response to nCRT. Analysis of DDR-related proteins by Luminex xMAP. The data is presented as % MFI for each protein marker per patient for the indicated response groups. **Independent set 1 (n=48) (A-G).** Here, **(A)** total ATR, **(B)** Chk1^S345^, **(C)** Chk2^T68^, **(D)** γH2AX^S139^, **(E)** p53^S15^, **(F)** total MDM2, and **(G)** total p21. **(H)** Associations among the protein markers assessed by xMAP and the foci-based assay for γH2AX^S139^. All markers, excluding p21, strongly correlated with each other. The color gradient represents the degree of positive (blue) or negative (red) correlation. **Independent set 2 (n=44) (I-O).** Here, **(I)** total ATR, **(J)** Chk1^S345^, **(K)** Chk2^T68^, **(L)** γH2AX^S139^, **(M)** p53^S15^, **(N)** total MDM2, and **(O)** total p21. Each black circle represents a patient. *p < 0.05, **p < 0.01, ***p < 0.001, NS (non-significant), using Kruskal-Wallis tests with by pairwise Wilcoxon rank sum tests adjusted for pre/post and time from sampling. The experiment was performed in duplicates and data from two independent experiments are presented.

We next tested the degree of association among the seven DDR proteins in the xMAP assay and found that each protein was strongly associated with all the others except total p21 (**Figure 3H**, Spearman’s correlation). Similarly, when comparing the data from the γH2AX^S139^ foci-based assay against the seven proteins in the DDR xMAP assay, we found a strong association with each except total p21 (**Figure 3H**, Spearman’s correlation). **Supplementary figure 5A** shows statistically significant agreement between the γH2AX^S139^ results from each assay, positive γH2AX^S139^ foci as detected by immune fluorescence and protein levels measured by DDR xMAP (Pearson correlation, P=0.0083).

We then analyzed PBMCs of LARC patients from the second independent set that contained only two nCRT response groups (n=44, **Table 1**, CR and PoR groups). At baseline, the CR group had significantly elevated expression of total ATR, Chk1^S345^, and Chk2^T68^, activation of γH2AX^S139^, checkpoint-associated activation of p53^S15^, and total MDM2 relative to the PoR group (**Figure 3I-O**; all p-values <= 1e-5). The levels of another DDR-related protein, total p21, did not correlate with response status (**Figure 3O**).

Finally, univariate CART identified the thresholds for six of the DDR proteins that distinguished the PoR and CR groups of patients (**Figures 4A-F**; all p values < 0. 001). No statistically significant cut-point was obtained for total p21 **(Supplementary figure 5B). Figure 4G** summarizes the study’s findings, demonstrating that the xMAP assay, performed on PBMCs isolated from blood specimens, effectively predicts CRs and PoRs among LARC patients in the population. These data suggest its potential utility in stratifying patients for nCRT treatment.

**Figure 4.**
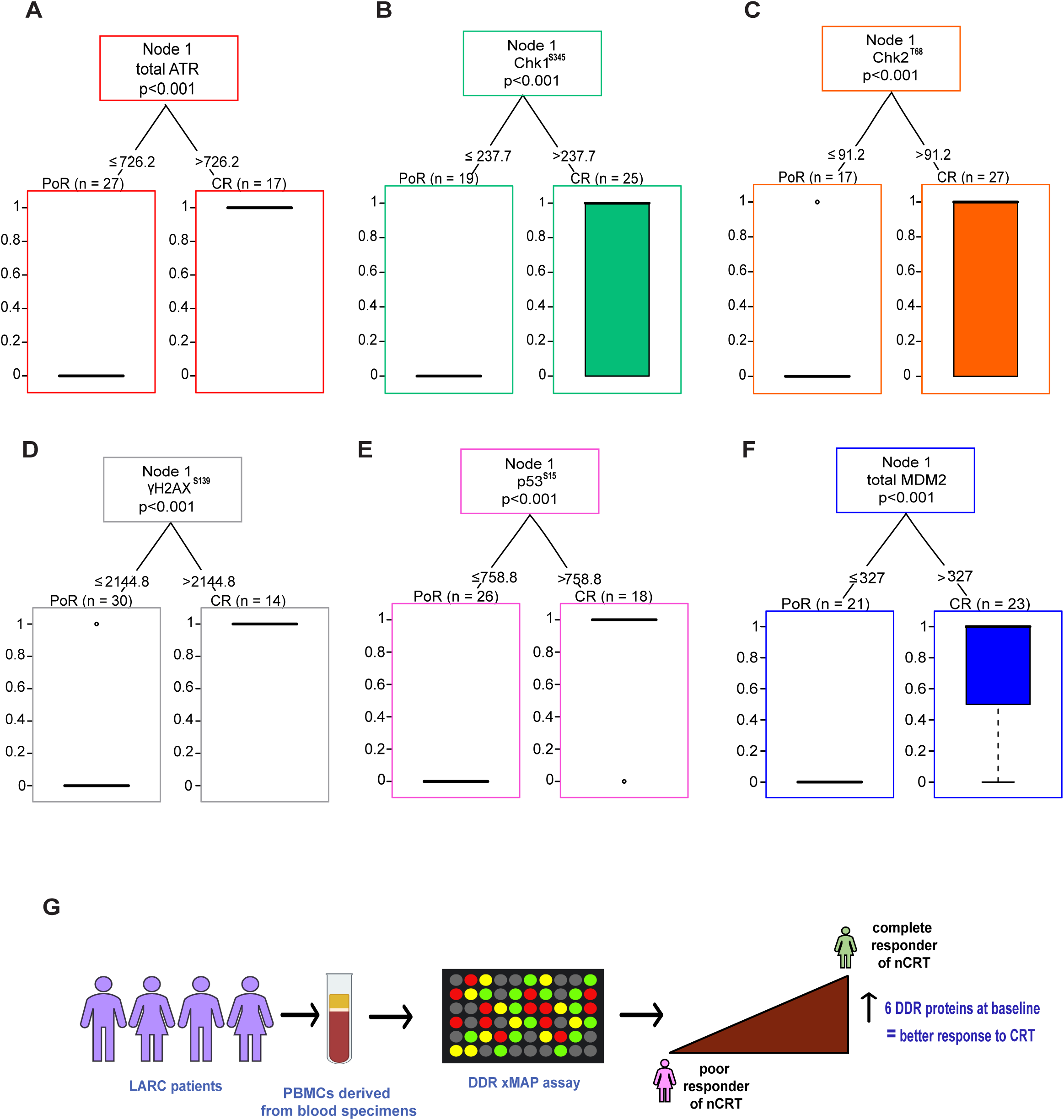
Univariate CART approach showing significant split in the levels of six DDR proteins and patient response. Significant splits were observed for the levels of six DDR-related protein markers, **(A)** total ATR, **(B)** Chk1^S345^, **(C)** Chk2^T68^, **(D)** γH2AX^S139^, **(E)** p53^S15^, and **(F)** total MDM2 that delineates patient response. The 1^st^ node represents the total number of cases with the branches that lead to a specific binary partition based on the optimal cut-points identified by the method, for poor response and complete response. Nodes 2 and 3 in each figure contain box-plots representing the distributions of poor (coded as 0) and complete responses (coded as 1) as determined by CART. Scenarios where all patients were classified correctly into their response group result in horizontal lines at 0 or 1. **(G)** Summary of the findings, demonstrating that the xMAP assay can distinguish complete responders from poor responders among LARC patients by detecting increased baseline expression of DDR proteins in PBMCs derived from blood specimens.

## Discussion

This study addresses the critical need to stratify patients effectively for nCRT response in LARC patients by identifying key DDR proteins and optimizing and validating an assay to accurately measure them and predict treatment benefit. By focusing on systemic DDR indicators in PBMCs rather than tumor markers, this method potentially offers a more accessible, less invasive predictor of treatment response. The elevated basal levels of DNA damage in PBMCs suggest that patients who achieve a CR following treatment may have underlying defects in DNA repair pathways, particularly in their ability to repair DSBs. This deficiency could make CR patients more susceptible to the accumulation of DNA damage, thereby enhancing the efficacy of DNA-damaging therapies such as radiation. In contrast, patients with PoR/PR may have more efficient DNA repair mechanisms, allowing them to better manage treatment-induced damage and reducing treatment efficacy. The susceptibility of CR patients’ PBMCs to DNA damage, both in basal conditions and after ionizing radiation, supports this hypothesis of compromised DNA repair. Further, the heightened sensitivity of lymphoblastoid cell lines from CR patients to DNA-damaging therapies points to a systemic defect in DDR capacity, potentially involving key DSB pathways such as homologous recombination. Overall, this approach could lead to more personalized, effective treatment regimens for LARC patients by identifying those likely to benefit from nCRT.

Currently, no definitive biomarkers predict the response to nCRT in LARC patients (5–8). Numerous studies have explored biomarkers to predict therapy response in LARC patients, primarily analyzing tumor mutations, gene expression profiles, and tumor microenvironment infiltrates (21–24). Notably, no somatic mutations have definitively correlated with response to nCRT. Chow et al. showed that *KRAS* and *TP53* mutations in LARC are independently associated with decreased response to nCRT (reduced pCR) and with lymph node metastasis (22), though these findings have yet to be definitively validated in subsequent studies. Similarly, transcriptomic analyses (21,24) and circulating tumor DNA (49) have failed to yield any consistent predictors of nCRT response. These challenges underscore the urgent need for reliable, minimally invasive biomarkers that can effectively guide treatment decisions and improve prognosis by accurately predicting therapeutic outcomes. The existing methods of clinical assessment are incapable of reliably predicting response to nCRT. Current management decisions in LARC are made based on the side effect profile of each treatment, the risk of locoregional or distant recurrence, the practice patterns of the clinicians, and the convenience of the patient (50). However, this does not consider the likelihood of response to therapy-a critical gap in our current treatment tailoring capabilities that emphasizes the potential benefits of biomarkers for LARC management.

Prior research consistently highlights inherent variations in DNA repair capacity among individuals, which plays a crucial role in cellular response to cancer therapy. We have shown that lymphocytes from patients with genetically unexplained early-onset and/or familial colorectal cancer have increased DNA DSBs compared to healthy cancer-free controls (35). Moreover, lymphocytes from endometrial cancer patients showed high baseline DNA damage and reduced repair capacity versus healthy controls (51). However, non-small cell lung cancer patients with enhanced DNA repair capacity exhibited lower survival rates in their peripheral lymphocytes (52). Supportively, studies in breast cancer patients have suggested that monitoring DNA damage in lymphocytes could assess the biological effects of DNA-damaging therapy and associated cancer risks (53,54). Additionally, EBV-transformed lymphoblastoid cell lines from BRCA2+/- carriers show increased DSB damage and reduced DSB repair, emphasizing the significant impact of haploinsufficiency on defective DSB repair (55). Collectively, variation in DNA repair capacity is important for predicting and improving individual responses to cancer treatment.

Historically, technologies like γH2AX^S139^ foci counting or the comet assay have been explored to reliably detect DDR in human biospecimens (12,35,36,45–48). However, these methods have suffered from significant inter-lab variability, which has limited their clinical application. In response to these challenges, we optimized and validated a Luminex-based xMAP assay for activated proteins during DDR, leveraging the powerful and versatile Luminex platform used in clinical and research applications. This study represents a significant advancement in approaches for patient stratification in LARC, offering a robust and predictive tool that addresses a longstanding gap in the field.

Finally, several limitations must be considered. Although our findings are significant, larger and more diverse cohorts are needed to confirm the generalizability across different populations and settings. As a future step, we aim to comprehensively analyze the inherent biological variability among patients, including genetic mutations, polymorphisms, and epigenetic changes, enhancing the predictive power. The results presented here require multi-center studies to ensure broader applicability. Additionally, this study primarily assessed protein levels in relation to immediate nCRT response; overall survival and disease-free survival were not evaluated. Future research should investigate how systemic DDR capabilities correlate with long-term outcomes. Future research should also consider incorporating specimens from the recent new treatment paradigms for patients with LARC (4,56–60). These include effective chemotherapy regimens, better approaches for radiotherapy, and the optimal sequence of these modalities in LARC and present new treatment strategies for LARC patients (4,56–60). These new options allow treatment choices to be made based on side effect profiles for appropriate patients; however, there are no available methods for predicting response to the various treatment options as a part of the selection process.

Overall, our study demonstrates that a blood-based systemic DDR signaling assay can effectively predict nCRT response in LARC patients. This assay could facilitate personalized treatment strategies, improving patient outcomes and optimizing therapeutic approaches in the evolving landscape of LARC management. Further research and validation are essential to fully realize the potential of this approach in clinical practice.

## Conclusion

We optimized and validated an xMAP assay to assess DDR signaling proteins in human PBMCs and identified that systemic DDR capabilities can predict response to nCRT in patients with LARC. This approach offers a more personalized method for treatment recommendations than standard anatomic staging. The differences in DDR activity identified in this study provide a foundation for validation in larger cohorts from LARC patients and eventually as an investigational assay/biomarker in clinical studies.

## Acknowledgments

The work in this grant was supported by the resources and expertise provided by the FCCC Cell Culture, Biosample Repository, Biostatistics, Population Sciences, and High Throughput Screening Facilities. Patient samples used in this study were obtained under the FCCC BRF Institutional Review Board-approved protocol. Consent and authorization for the use of de-identified specimens and associated clinical data for research was obtained from all BRF participants prior to specimen collection. The BRF is supported by the FCCC Comprehensive Cancer Center Support Grant (CCSG) awarded by the National Cancer Institute (NCI P30 CA006927).

We also acknowledge the FCCC Summer Undergraduate Research Fellowship program and the Department of Radiation Oncology. We acknowledge Amanda Browne (formerly at Cancer Prevention and Control Program, FCCC), and Shreya Shah (formerly at Temple University, Philadelphia) for valuable assistance on the study. We also acknowledge the mentorship provided to E.V.D. by Dr. Ilya Serebriiskii at FCCC. Finally, I (S.A.) express immense gratitude to Dr. Margie Clapper (Professor and Co-Leader, Cancer Prevention and Control Program, FCCC) for her mentorship on the DOD Career Development Award, her time, and advice for my career development.

## Abbreviations

BRF: Biosample Repository Facility
CART: Classification and Regression Trees
CR: complete responders
nCRT: neoadjuvant chemoradiotherapy
CV: coefficient of variation
D: difference
DDR: DNA damage response
DSB: double stranded breaks
EBV: Epstein-Barr virus
FCCC: Fox Chase Cancer Center
IF: immunofluorescence
HTSF: High Throughput Screening Facility
IR: irradiation
IRB: institutional review board
OR: odds ratios
LARC: locally advanced rectal cancer
MFI: mean fluorescence intensity
NAR score: Neoadjuvant rectal score
PBMCs: peripheral blood monocytes
pCR: pathological complete response
PHA: phytohemagglutinin
PR: partial responders
PoR: poor responders
RT: room temperature
SAPE: Streptavidin-Phycoerythrin
SD: standard deviation
TNT: total neoadjuvant therapy
xMAP: multi-analyte platform.

## Author contributions

E.V.D, P.W.C, and A.H., performed most studies, data analysis, writing; V.A., clinical data abstraction, biospecimen selection, R.W.L., γH2AX^S139^ experiments, data analysis; E.H., P.W.C., and K.D., statistical analysis; J.D.J, B.M.S, and D.C.C., assistance with biospecimen selection and provision of specimens and clinical data; M.B.E., Luminex xMAP, γH2AX^S139^ microscopy (image acquisition, image export and analysis), experimental planning and analysis; D.B., planning Luminex experiments, discussion; E.A.G., discussion & writing; J.E.M & S.A., discussion, designing, planning studies, writing and funding.

