## Supplementary document for "Optimizing and Validating Systemic DNA Damage Response Profiling to Predict Neoadjuvant Chemoradiation Response in Rectal Cancer"

**Supplementary Methods.**

**Preparation of protein lysates.** Protein lysates were prepared directly from banked PBMCs (n=48) or banked-cultured PBMCs (n=50). For banked-cultured PBMCs, the banked PBMCs were cultured as described previously (37). The banked PBMCs (cryopreserved in 90% FBS and 10% DMSO) were received from the FCCC BRF and stored in -80 °C until lysates were prepared. For lysate preparation, samples were retrieved from -80 °C or liquid nitrogen and kept on dry ice. The water bath was set at 37 °C to thaw the sample in the bath for 2-4 minutes. Then 9 ml of FBS was added to the thawed samples to dilute the DMSO, and samples were spun for 10 minutes at 1300 rpm at 4 °C. FBS was discarded, and cells were washed with 1x PBS by re-spinning at 1300 rpm at 4 °C. The supernatant was discarded, and cells were placed on ice. Cell lysis buffer was freshly prepared by adding protease inhibitor cocktail Set III (1:100; 535140, EMD Millipore Sigma, Burlington, MA) to Milliplex Lysis Buffer (1x, 43-040, EMD Millipore Sigma, Burlington, MA). Further, 0.6 ml of the lysis buffer (ice cold) was added to each PBMC sample, mixed and transferred to cold 1.5 mL Eppendorf tubes. Samples were briefly vortexed and were rocked at 4 °C for 15 minutes on a shaker. Supernatants were collected by centrifugation at 15,000 rpm for 15 minutes at 4 °C. Protein content was quantified using Qubit™ Protein BR assay kit (A50668, Thermo Fisher Scientific, Waltham, MA) using the Qubit Fluorometer.

**xMAP Immunoassay protocol.** The assay was performed using the MILLIPLEX® xMAP assay kit (48-621MAG, Millipore Sigma, Burlington, MA) and the manufacturers protocol was followed.

**Vendor-provided xMAP assay controls.** Jurkat Cell Lysate: anisomycin (Cat. No. 47-207) was provided as a lyophilized stock of cell lysate prepared from Jurkat cells treated with 25μM anisomycin (4h); it is used as a stimulated control. A549 Cell Lysate: camptothecin (Cat. No. 47-218) was provided as a lyophilized stock of cell lysate prepared from A549 cells stimulated with 5μM camptothecin (overnight); it is also used as a stimulated control. HeLa Cell Lysate: lambda phosphatase (Cat. No. 47-229) was provided as a lyophilized stock of cell lysate prepared from unstimulated HeLa cells treated with lambda phosphatase; it is used as an unstimulated control. A549 (camptothecin-treated), Jurkat (anisomycin-treated) and HeLa (lambda phosphatase-treated) controls are vendor-supplied in the MILLIPLEX® MAP 7-plex DNA Damage/Genotoxicity Magnetic Bead Kit (Cat. No. 48-621MAG. Controls were diluted in MILLIPLEX® Assay Buffer to generate a semi-quantitative curve (1 mg/mL,0.5 mg/mL, 0.25 mg/mL, 0.125 mg/mL, 0.0627 mg/mL, 0.0312 mg/mL, and 0.0156 mg/mL).

**Preparation of xMAP magnetic beads.** MILLIPLEX® magnetic beads are provided as a 20x stock solution and should be protected from light. 20x stock magnetic beads were vortexed for 30 seconds. The beads were diluted to 1x by combining 0.150 mL beads with 2.85 mL of MILLIPLEX® Assay Buffer 1 in the mixing bottle provided with the kit. 1x magnetic beads were vortexed for 15 seconds. Before use, 1x beads were transferred in a reservoir using a pipette.

**Preparation of Biotin-Labeled Detection Antibody and Streptavidin-PE.** MILLIPLEX® Detection Antibody is provided as a 20x stock solution. 20x detection antibody was vortexed for 10 seconds and diluted to 1x by adding 2.85 mL of MILLIPLEX® Assay Buffer 1 in the mixing bottle provided with the kit. For the preparation of the MILLIPLEX® Streptavidin-Phycoerythrin 1:25 (SAPE), it was vortexed for 10 seconds. SAPE was diluted by combining 0.120 mL of Streptavidin-Phycoerythrin with 2.88 mL of MILLIPLEX® Assay Buffer 1 using one of the mixing bottles provided. 1x biotinylated detection antibody and SAPE were transferred with a pipette to separate reservoirs.

The reagents were allowed to reach room temperature prior to use. Protein lysates (5µg) were diluted 1:1 in MILLIPLEX® Assay Buffer. For all wells of the 96 well assay plate, 50 µL of Assay Buffer was added and the plate was sealed and agitated on a plate shaker (600-800 rpm) for 10 minutes at room temperature (20-25 °C). Assay buffer was removed, and the residual amount of the buffer was removed by inverting the plate and tapping it onto absorbent towels several times. To designated wells 25 µl of Assay Buffer (blank), 25 µl of vendor-provided controls, or 25 µl of protein lysate samples was added. To all wells 25 µL of 1x bead suspension was added. The plate was covered with a foil seal and incubated overnight (16-20h) at 2-8 °C on an orbital plate shaker (600-800 rpm) protected from light.

After the overnight incubation, the plate and shaker were removed from 2-8 °C and the plate was agitated as before for 30 minutes at room temperature prior to proceeding. The 96-well magnetic separation block (Cat. No. 40-285) was attached to the plate for 60 seconds to collect the beads before decanting the samples and controls. The samples are decanted in the sink and pressed on a paper towel while attached to the magnetic separation block. The plate was removed from the magnetic separation block and washed twice with 100 µL Assay Buffer per well placing the 96-well plate back on the shaker after the addition. For each wash, the plate was attached to the magnetic separation block and beads are collected via magnetic separation prior to decanting the Assay Buffer. After washes and discarding the supernatant, 25 µL of 1x MILLIPLEX® Detection Antibody was added to all wells. The plate was sealed and incubated with agitation on the plate shaker for 1h at room temperature (20-25 °C). Beads were collected via magnetic separation as before, and the Detection Antibody was decanted. After the removal of the Detection Antibody, 25 µL of 1X MILLIPLEX® Streptavidin-Phycoerythrin (SAPE) was added to all wells. The plate was sealed and incubated with agitation for 15 minutes at room temperature (20-25 °C).

After this incubation, 25 µL of MILLIPLEX® Cell Signaling Amplification Buffer was added to each well (note: do not remove SAPE). The plate was sealed and incubated with agitation for 15 minutes at room temperature (20-25 °C). Beads are collected via magnetic separation as before, and the SAPE/Amplification buffer was removed. The beads were suspended in 150 µL of MILLIPLEX® Assay Buffer, mixed as before on a plate shaker for 5 minutes, and the samples were analyzed using a Bioplex 200.

**Supplemental Figure Legends.**

**Supplementary figure 1. (A) γH2AX$^{S139}$ foci detected in pre- or post-treatment PBMCs of matched rectal cancer patients.** PBMCs from matched patients (n=11) were quantified for γH2AX$^{S139}$ foci and the data is represented as the percent positive γH2AX$^{S139}$ foci, normalized to pre-CRT sample per patient. *p=0.4961 (Wilcoxon signed-rank test), using logistic regression model adjusted for age, gender, time from treatment. Black circle represents pre-CRT and white circle- pre-Surgery sample for each patient. **(B) Correlation of γH2AX$^{S139}$ foci detected in PBMCs of LARC patients with age.** Graph represents association between γH2AX$^{S139}$ positive foci data from PBMCs and the age of the study cases (n=54, independent set 1). Association was tested using Spearman's rank correlation, P=0.8315.

**Supplementary figure 2. xMAP standardization with assay controls**. Graphs show development of a semi-quantitative curve using assay controls. Data for all seven markers, **(A)** total ATR, **(B)** Chk1$^{S345}$, **(C)** Chk2$^{T68}$, **(D)** γH2AX$^{S139}$, **(E)** p53$^{S15}$, **(F)** total MDM2, and **(G)** total p21, is shown from 12 technical replicates with standard deviation.

**Supplementary figure 3. Detailed assessments of individual seven DDR xMAP assay markers from healthy control study participants with calibration curves of the assay control 1.** The data shown are from lysates prepared from banked PBMCs and banked-cultured PBMCs of healthy controls. **From banked PBMCs, Top (A-E). Assay control 1: Jurkat cells treated with anisomycin.** The red squares represent the semi-quantitative curve for the assay control 1, and each blue circle represents data from an individual healthy control (n=48). The dotted lines represent the standard deviation along the solid line. Replicates are shown from 12 replicates each run as duplicates. Concentration is in mg/ml. **From banked-cultured PBMCs, Bottom (F-J). Assay control 1: Jurkat cells treated with anisomycin.** The red squares represent the semi-quantitative curve for the assay control 1, and each blue circle represents data from an individual healthy control (n=50). The dotted lines represent the standard deviation along the solid line. Replicates are shown from 12 replicates each run as duplicates. Concentration is in mg/ml.

**Supplementary figure 4. Data showing details of assessments of individual seven DDR xMAP assay markers from healthy control study participants with calibration curves of the assay control 2.** The data shown are from lysates prepared from banked PBMCs and banked-cultured PBMCs of healthy controls. **From banked PBMCs, Top (A-F). Assay control 2: A549 cells treated with camptothecin.** The red squares represent the semi-quantitative curve for the assay control 1, and each blue circle represents data from an individual healthy control (n=48). The dotted lines represent the standard deviation along the solid line. Replicates are shown from 12 replicates each run as duplicates. Concentration is in mg/ml. **From banked-cultured PBMCs, Bottom (G-L). Assay control 2: A549 cells treated with camptothecin.** The red squares represent the semi-quantitative curve for the assay control 1, and each blue circle represents data from an individual healthy control (n=50). The dotted lines represent the standard deviation along the solid line. Replicates are shown from 12 replicates each run as duplicates. Concentration is in mg/ml.

**Supplementary figure 5. (A) Association of γH2AX$^{S139}$ findings using the two detection techniques.** γH2AX$^{S139}$ results obtained using the IF method (x-axis) compared to results obtained using xMAP (y-axis). The observed correlation per response group is statistically significant, P<0.001. For correlation analysis, Pearson nonparametric correlation coefficient with two-tailed p-value was used. **(B) Univariate CART analysis.** Result from univariate CART analysis shows no significant split in the levels of total p21 that could delineate patient response in LARC patient biospecimens.

**Supplementary Table Legends.**

**Supplementary table 1.** Coefficient of determination ($R^2$) for DDR markers for vendor-provided assay controls 1 and 2 showing the goodness of fit.

**Supplementary table 2.** Demographic characteristics of healthy controls.

**Supplementary table 1.** Coefficient of determination ($R^2$) for DDR markers for vendor-provided assay controls 1 and 2 showing the goodness of fit.

| Assay control 1 | | | | | | | |
|---|---|---|---|---|---|---|---|
| **Marker** | **total ATR** | **CHK1$^{S345}$** | **CHK2$^{T68}$** | **γH2AX$^{S139}$** | **p53$^{S15}$** | **total MDM2** | **total p21** |
| **Goodness of Fit** | | | | | | | |
| Degrees of Freedom | 2 | 3 | 3 | 2 | 1 | 3 | 1 |
| R squared | 0.9978 | 0.9997 | 1 | 0.9994 | 0.9357 | 0.9999 | 0.9836 |
| Adjusted R squared | 0.9945 | 0.9995 | 1 | 0.9986 | 0.743 | 0.9999 | 0.9345 |
| Sum of Squares | 19.84 | 27.12 | 11.02 | 1280 | 1.617 | 56.31 | 5.064 |
| **Assay control 2** | | | | | | | |
| **Marker** | **total ATR** | **CHK1$^{S345}$** | **CHK2$^{T68}$** | **γH2AX$^{S139}$** | **p53$^{S15}$** | **total MDM2** | **total p21** |
| **Goodness of Fit** | | | | | | | |
| Degrees of Freedom | 3 | 3 | 3 | 3 | 3 | 3 | 2 |
| R squared | 0.9947 | 0.9798 | 0.9984 | 0.9988 | 0.9991 | 0.9999 | 0.9967 |
| Adjusted R squared | 0.9894 | 0.9596 | 0.9968 | 0.9976 | 0.9983 | 0.9998 | 0.9917 |
| Sum of Squares | 42.84 | 3663 | 32.13 | 51.37 | 259.8 | 750 | 589606 |

**Supplementary table 2.** Demographic characteristics of healthy controls.

| **Characteristics** | |
|---|---|
| **Age (years)** | |
| Mean | 49 |
| Median | 49 |
| Range | 32-71 |
| **Gender, total N (%)** | |
| Male | 18 (36) |
| Female | 32 (64) |
| **Self-reported Race, total N (%)** | |
| Black | 10 (20) |
| White | 37 (74) |
| More than one race | 3 (6) |

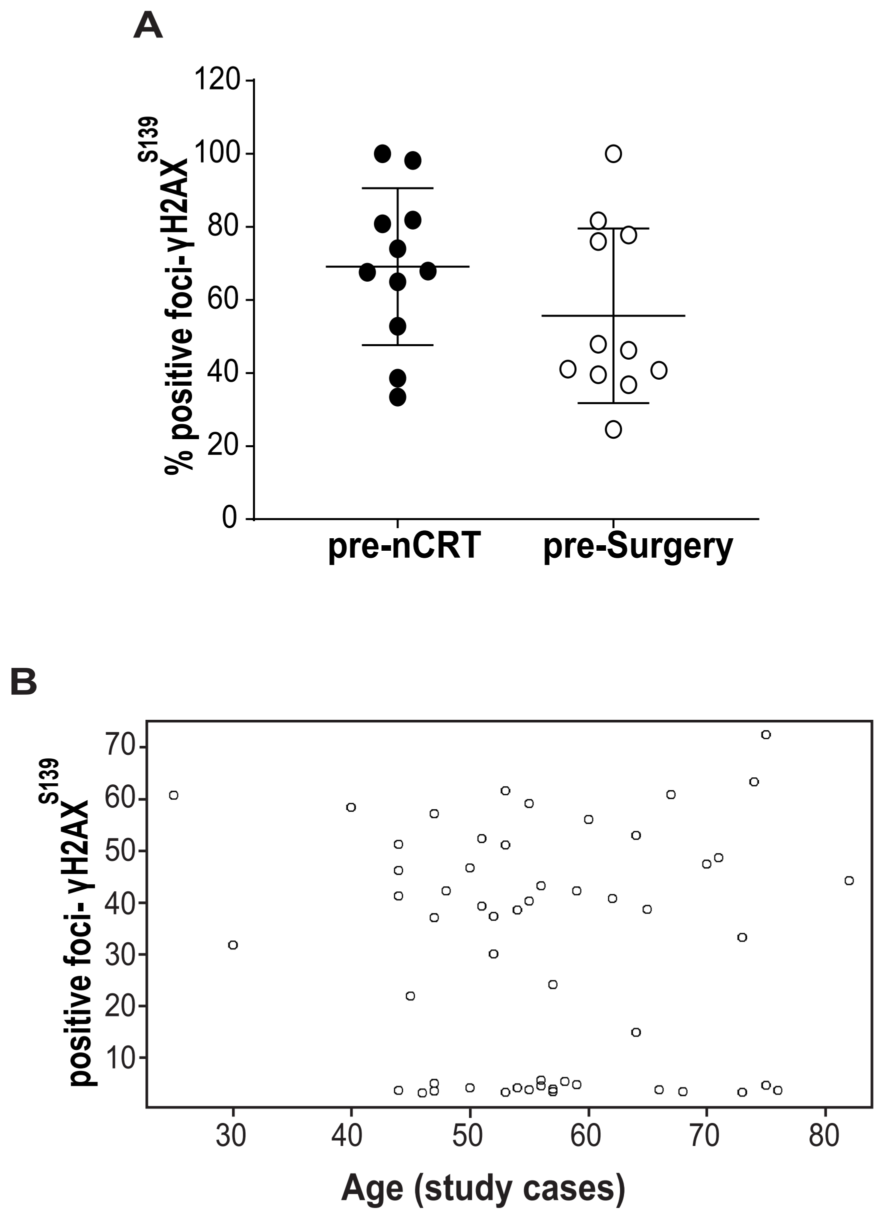

**Supplementary figure 1. Demidova, Czyzewicz, and Hasan et al.**

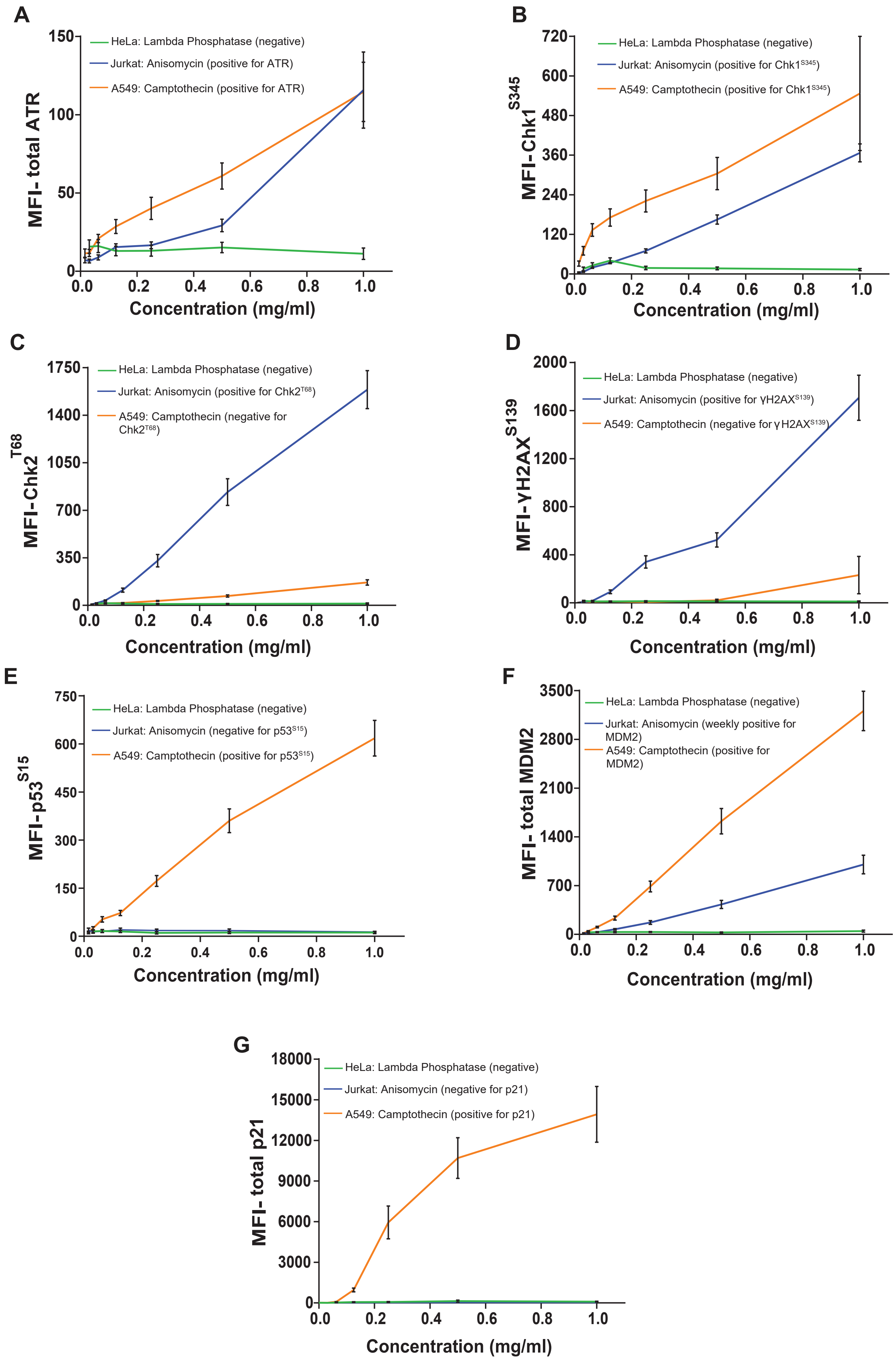

**Supplementary figure 2. Demidova, Czyzewicz, and Hasan et al.**

### Assay control 1: Jurkat (treated with Anisomycin) banked PBMCs

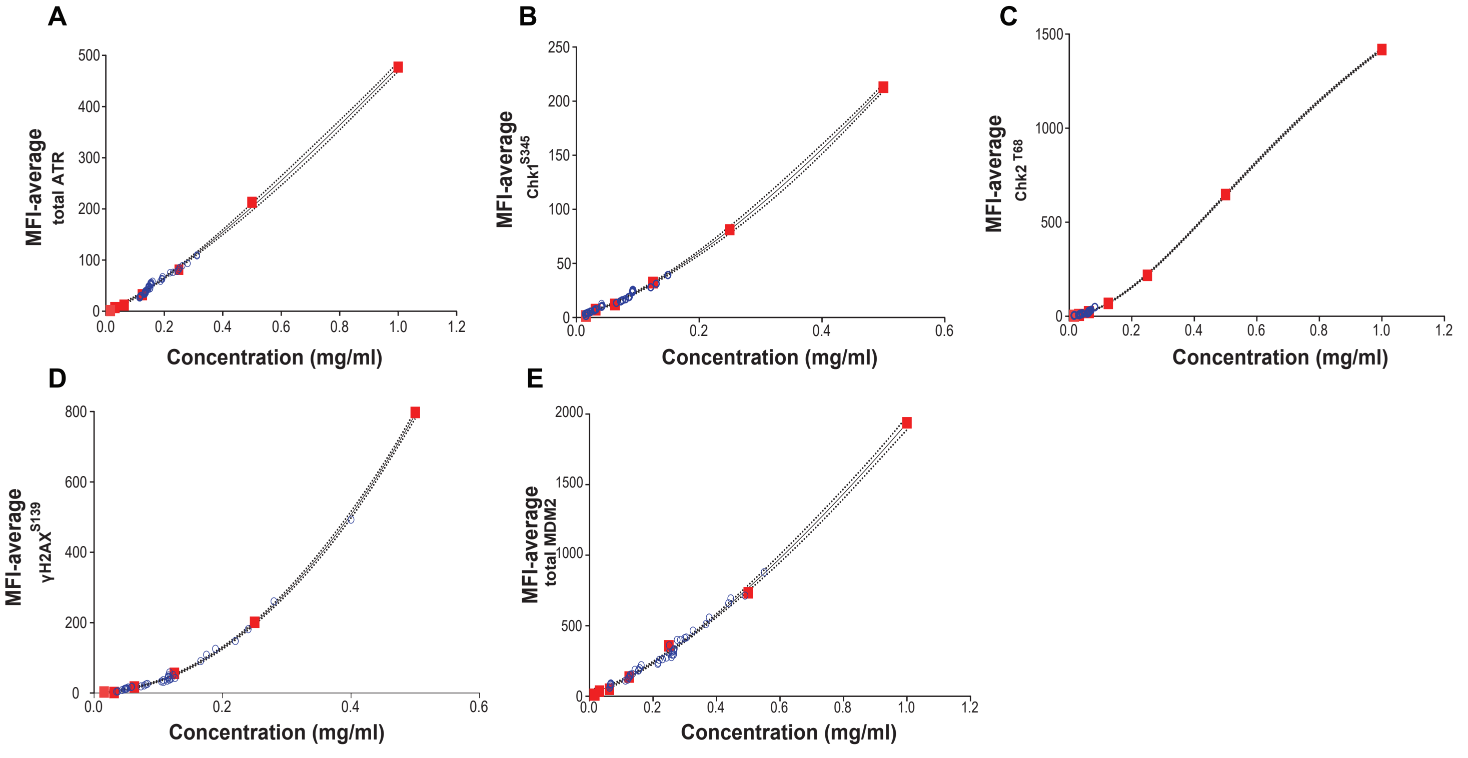

### Assay control 1: Jurkat (treated with Anisomycin) banked-cultured PBMCs

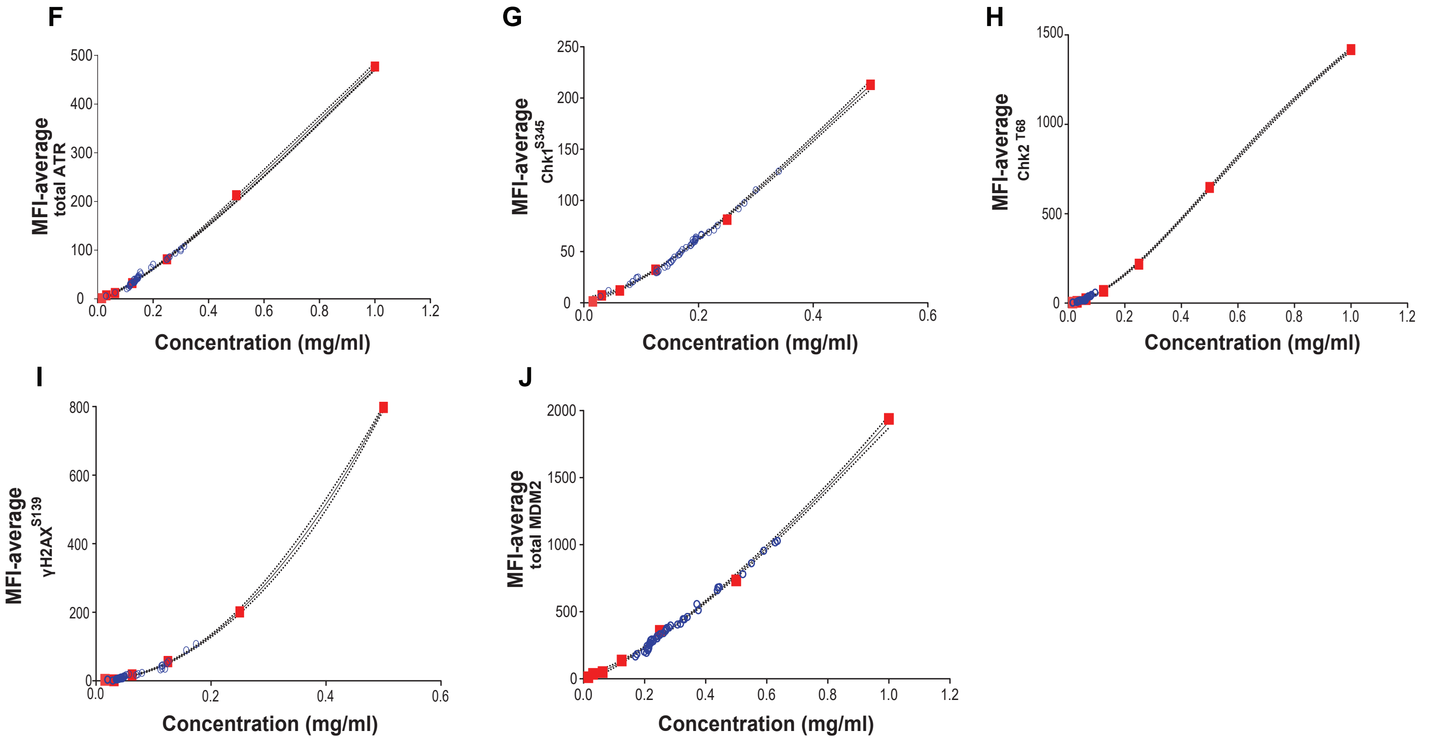

Supplementary figure 3. Demidova, Czyzewicz, and Hasan et al.

### Assay control 2: A549 (treated with Camptothecin)
### banked PBMCs

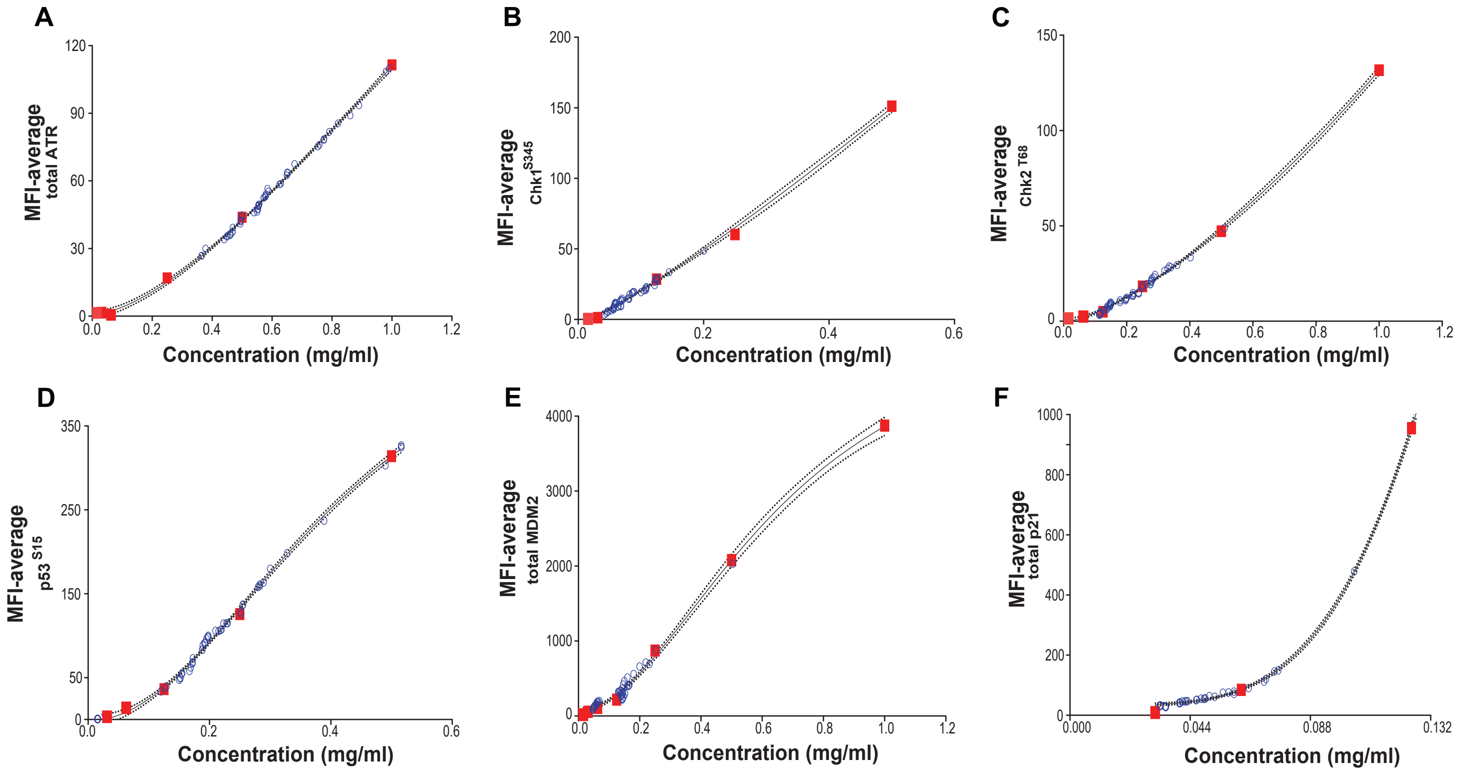

### Assay control 2: A549 (treated with Camptothecin)
### banked-cultured PBMCs

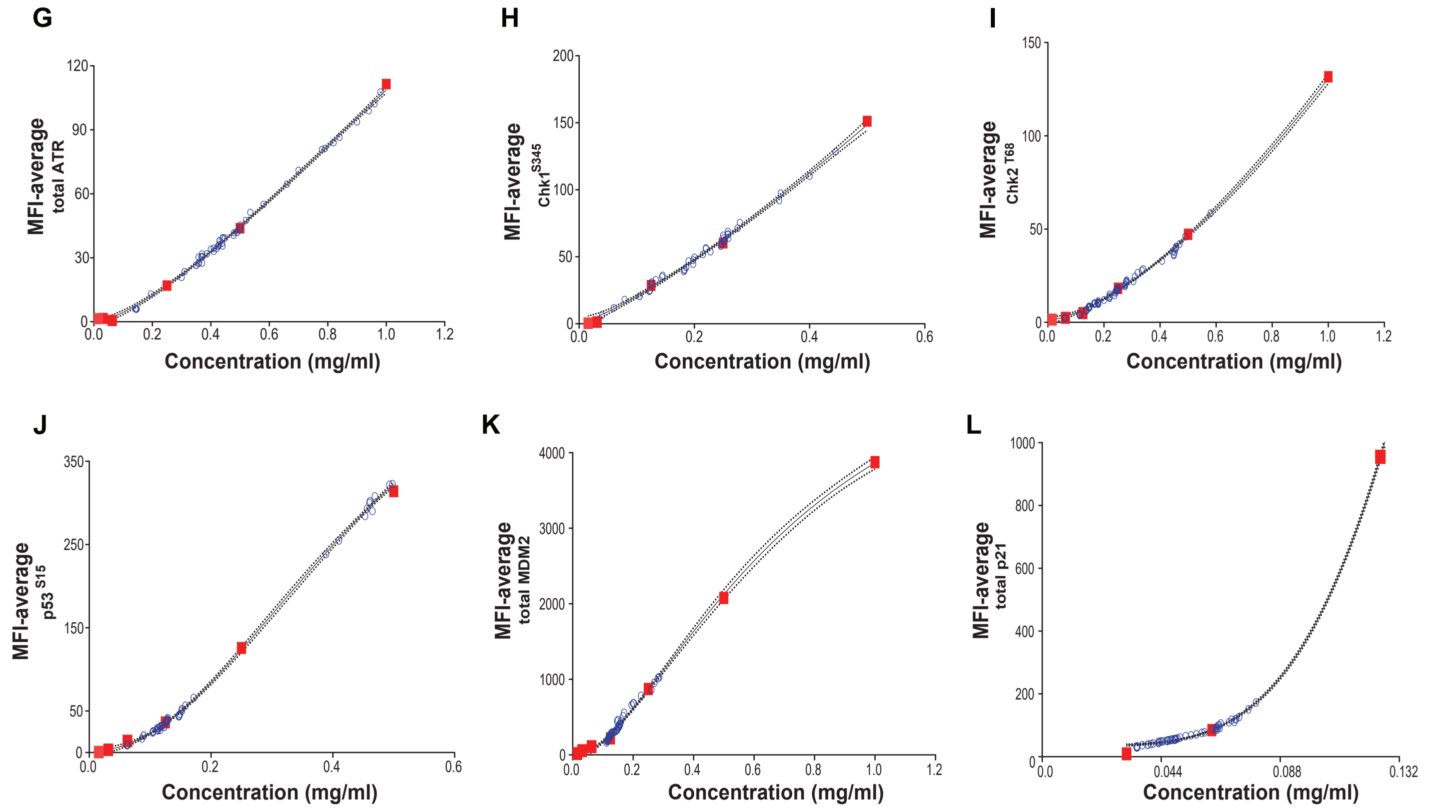

**Supplementary figure 4. Demidova, Czyzewicz, and Hasan et al.**

A

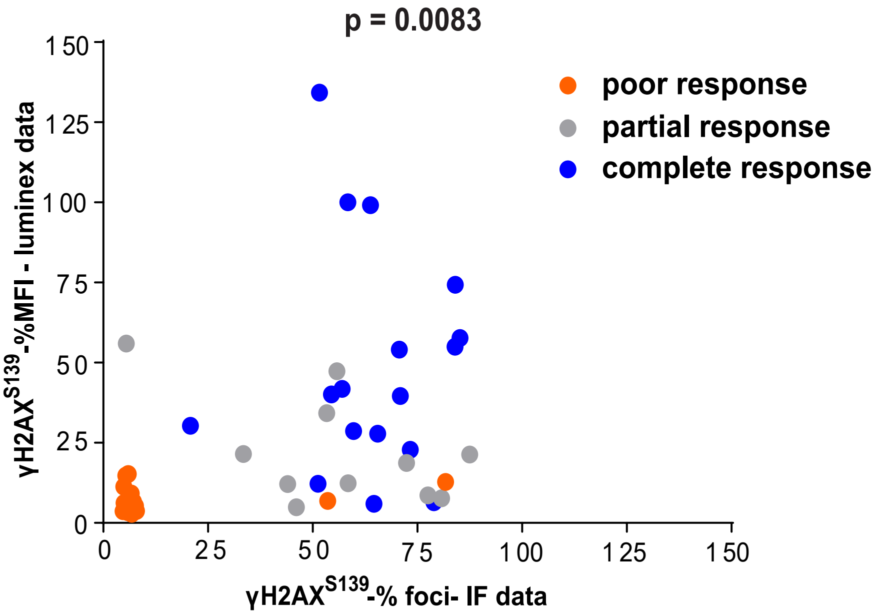

B

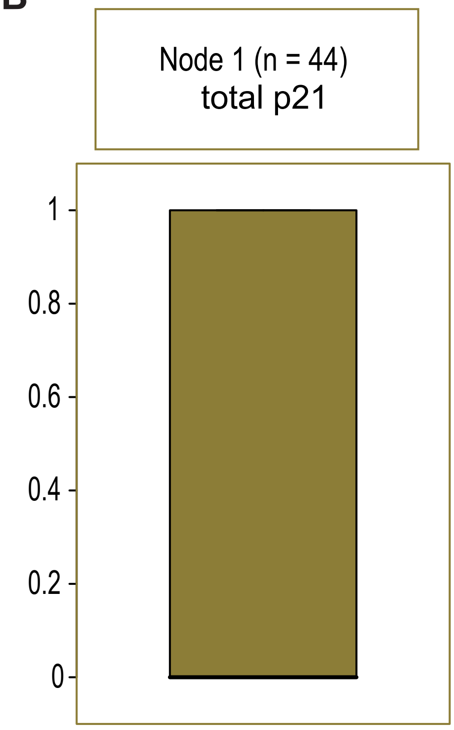

Supplementary figure 5. Demidova, Czyzewicz, and Hasan et al.
